# High agreement between lean mass assessed by Dual-Energy X-ray absorptiometry and Bioelectrical Impedance Analysis across the adult lifespan

**DOI:** 10.1101/2025.02.03.25321573

**Authors:** T. Kristensen, R.S. Kamper, B. Haddock, H. Nygaard, T. Nørst, P. Hovind, C. Suetta

**Affiliations:** Department of Clinical Physiology, Nuclear Medicine and PET, Rigshospitalet Glostrup, Glostrup, Denmark; Geriatric Research Unit, Department of Geriatric and Palliative Medicine, Copenhagen University Hospital, Bispebjerg and Frederiksberg, Copenhagen, Denmark; Department of Emergency Medicine, Copenhagen University Hospital, Bispebjerg and Frederiksberg, Copenhagen, Denmark; CopenAge, Copenhagen Center for Clinical Age Research, University of Copenhagen, Copenhagen, Denmark; Department of Clinical Physiology and Nuclear Medicine, Copenhagen University Hospital, Bispebjerg and Frederiksberg, Copenhagen, Denmark

**Keywords:** Bioelectrical impedance analysis, BIA, DXA, Dual-energy X-ray absorptiometry, appendicular lean mass, sarcopenia

## Abstract

**Background:** Bioelectrical impedance analysis (BIA) is considered a portable alternative to Dual-energy X-ray absorptiometry (DXA) when assessing deficient muscle mass, a major concern for older adults associated with increased risks of falls, fractures, disability, and mortality. However, several studies have demonstrated that BIA overestimates muscle mass in the older population and that estimates of muscle mass may depend on the BIA and DXA devices utilized. This study aims to describe the concordance between lean mass assessed by DXA and BIA across the adult population.

**Methods:** This study is based on data from the Copenhagen Sarcopenia Study, a cross-sectional study of 1116 healthy participants aged 22-92 years with complete measurements of lean mass from both DXA (iDXA, GE Lunar) and BIA (Inbody 770). Relative appendicular lean mass (ALM) was calculated by normalizing ALM to height^2^ to account for allometric differences in body size.

**Results:** For the total cohort, linear regression models showed a strong correlation between BIA and DXA for ALM (R² = 0.95) and relative ALM (R² = 0.88). A small, yet consistent, bias of relative ALM values was observed where BIA measures were on average 0.25±0.50 kg/m^2^ higher compared to measurements by DXA. The bias was consistent across age groups with a mean difference in relative ALM of 0.24±0.50 kg/m^2^ and 0.26±0.49 kg/m^2^ for younger and older participants, respectively.

**Conclusion:** BIA is a reliable alternative to DXA for assessing lean mass, in both younger and older adults. The findings indicate that with simple correction equations for ALM and relative ALM, BIA can be a practical and accessible tool in clinical settings where DXA is unavailable. These equations make BIA particularly useful for diagnosing sarcopenia in a clinical setting, allowing for early detection and management regardless of age.

**Key messages:** Bioelectrical impedance analysis (BIA) is considered a portable alternative to Dual-energy X-ray absorptiometry (DXA) when assessing low muscle mass; a major concern for older adults which is associated with increased risks of falls, fractures, disability, and mortality. However, several studies have demonstrated that BIA overestimates muscle mass in the older population and estimates of muscle mass may depend on the BIA and DXA devices utilized. By applying the models of regression equations developed in this study, clinicians can improve the accuracy of BIA measurements, making BIA a practical alternative to DXA in clinical settings where DXA is not accessible. These findings are especially relevant for the early detection and management of sarcopenia in older adults, allowing for timely interventions to preserve muscle function and reduce the risk of adverse health outcomes.

## Introduction

Skeletal muscle mass is essential for maintaining mobility, metabolism, and overall health, particularly as individuals age [1,2]. The decline in muscle mass and muscle strength, known as sarcopenia, is a major concern for older adults and is associated with increased risks of falls, fractures, disability, and mortality [3–7]. Sarcopenia, either alone or in combination with other conditions, exacerbates the effects of chronic diseases such as cardiovascular disease, diabetes, and cancer [8–12]. Given the critical role of muscle mass in health outcomes, accurate assessment of lean body mass is vital for diagnosing sarcopenia and guiding interventions aimed at maintaining muscle health.

Muscle mass can be assessed using a variety of methods. Magnetic resonance imaging (MRI) and computed tomography (CT) are considered gold-standard methods due to the high resolution, accuracy and the ability to evaluate muscle architecture and presence of intramuscular fat [13,14]. However, both methods are time and labor demanding and have limited availability, due to high costs and radiation exposure (CT) limiting the use in daily clinical practice [15].

Dual-Energy X-ray absorptiometry (DXA) is another method for estimating lean mass and is the preferred choice for many researchers and clinicians [16]. DXA uses two different energy spectra and this assessment can differentiate total lean soft tissue from fat and bone mass based on the X-ray absorption [17]. As a result, DXA estimates both total lean mass and appendicular lean mass (ALM) [17]. ALM, which represents the lean mass of the limbs, is a strong predictor of physical function, and is commonly used to diagnose the lean mass component of sarcopenia [18]. However, despite its precision, DXA has several disadvantages limiting a more widespread use. DXA machines are expensive, require dedicated facilities and trained personnel, and are not easily portable. This makes the DXA impractical for routine clinical use or in remote settings.

Bioelectrical Impedance Analysis (BIA) is a popular alternative to DXA for assessing body composition and has been suggested as a more practical and affordable alternative [15,18–21]. The BIA devices are portable and simple to use, which makes them more accessible in clinical settings. Body composition is estimated based on the electrical conductivity of tissues, with lean tissues (muscles) conducting electricity better than fat tissue [16]. BIA does not provide a direct measure of lean mass but estimates both total and appendicular lean mass using equations based on the unique electrical conductivity properties of each tissue type [18]. The method is inexpensive, simple, and can be used in the assessment of muscle mass in both ambulatory and immobile patients [15].

While BIA offers practical advantages, studies have reported some BIA devices to overestimate muscle mass compared to DXA, particularly in older adults [22–24]. BIA accuracy can be influenced by the specific device used, as well as factors such as hydration status, fat distribution, and population characteristics [22–24].

Given the growing clinical demand for reliable muscle mass assessments, this study aims to compare the agreement between BIA and DXA for measuring ALM and relative ALM (normalized to height²) in a large and diverse adult population. By investigating potential sources of confounders, such as height, age, weight, sex, and BMI, the aim of this study is to identify the conditions, and possible corrections, necessary for the accurate implementation of ALM measured using the InBody 770 BIA device. This knowledge would greatly improve the clinical utility of BIA, particularly for diagnosing sarcopenia and monitoring muscle mass across the adult lifespan.

## Methods

### Subjects and study design

This study is a part of the Copenhagen Sarcopenia Study, a population-based cross-sectional investigation conducted at Copenhagen University Hospital, Rigshospitalet Glostrup, between December 2013 and June 2016 [25,26]. The current study comprises 1116 healthy adults (55% women and 45% men) aged between 22 and 92 years with complete measurements of lean mass from both DXA (iDXA, GE Lunar) and BIA (Inbody 770). In brief, inclusion criteria required participants to be generally healthy, without any chronic conditions known to affect body composition, such as severe obesity, heart failure, or neuromuscular disorders. Exclusion criteria included pregnancy, recent surgery, use of medications that affect fluid balance or muscle mass, and any condition preventing them from standing unassisted.

All participants provided written informed consent, and the study was approved by the Ethical Committee of the Capital Region and Frederiksberg (H-3-2013-124) and conducted in accordance with the Declaration of Helsinki II.

### Anthropometry

All measurements were performed by three trained health research employees. Height was measured without shoes to the nearest 0.1 cm and body weight was assessed wearing light clothing to the closest 0.1 kg. Body mass index (BMI) was calculated as weight/height^2^.

### Dual-Energy X-ray absorptiometry

Whole-body DXA scans were performed using a GE iDXA fan-beam scanner (GE Lunar, Madison, WI, USA), operated by one of three appointed and trained technicians to ensure consistency. The same device was used for all body scans and analyses of all exams were done using Encore software version 16.0. Lean soft tissue assessed by DXA is composed of all fat-free mass components except for mineral content. ALM was determined as the sum of lean soft tissue from the arms and legs. To account for differences in body size, relative ALM was calculated by dividing ALM by the square of height (ALM/h²).

### Bioelectrical impedance analysis

Bioimpedance analysis (BIA) was measured using the InBody 770, a multi-frequency Bioelectrical Impedance device (InBody Corp., Seoul, Korea). The device uses an eight-electrode system to send low-level electrical currents through the body. Participants were instructed to hold the electrodes while standing barefoot on the other electrodes on the device platform as per the manufacturer’s instructions. Appendicular lean mass (ALM) was calculated as the sum of the lean mass of the upper and lower extremities. To account for differences in body size, relative ALM was calculated by dividing ALM by the square of height (ALM/h²).

### Statistical analyses

All statistical analyses were performed using SPSS Statistics 25. Unless otherwise stated, data are presented as group mean ± standard deviation (SD). Analyses were performed for all participants as a single cohort, divided into young and old, and separately for women and men.

Relative appendicular lean mass was calculated by dividing lean mass values in kilograms by the square of height in meters (kg/m^2^). Statistical differences between women and men were evaluated using Student’s t-test for independent samples. In addition, the relationships between age, weight, height, and measured variables were assessed by regression analyses. Least square linear and quadratic regression models were assessed using the coefficient of determination (R^2^) to determine the most appropriate regression model. For the analysis of variance, residuals were checked for normal distribution by one-sample Kolmogorov-Smirnov tests and visual inspection.

Pearson correlations were performed to examine the relationship between ALM values measured by the BIA and the DXA. Agreement between methods was assessed using the Bland-Altman plot [27] and reliability using the Intra-Class Correlation Coefficient (ICC). We considered an ICC over 0.90 as very high, between 0.70 and 0.89 as high and between 0.50 and 0.69 as moderate [28]. Stepwise multiple regression was used to explore potential contributions of factors such as height, sex, age, weight, and BMI. Differences were considered significant at *P <* 0.05.

## Results

A total of 1116 participants (55% women and 45% men) were included in the analyses. The mean age of the participants was 57 ± 0.51 years, with a range of 22–92 years. Table 1 presents the baseline characteristics of the cohort. The height and weight of men were significantly higher than women, resulting in a higher mean BMI (all P < 0.001). Men also had significantly higher absolute and relative ALM values by both BIA and DXA measurements (all P < 0.001). All parameters for younger participants were significantly higher (all P < 0.001) than for the older participants except for body weight (P = 0.221).

**Table 1.** Characteristics of the study cohort.

|  | **Women**<br>**(n = 616)** | **Men**<br>**(n = 500)** | **p-**<br>**value** | **Old**<br>**(n = 442)** | **Young**<br>**(n = 674)** | **p-value** |
| --- | --- | --- | --- | --- | --- | --- |
| Age (years) | 58 (42; 71) | 59 (45; 71) | 0.819 | 74 (69; 77) | 47 (37; 56) | <0.001 |
| Weight (kg) | 66.0 (60.0; 74.0) | 82.0 (75.0; 91,4) | <0.001 | 73.0 (63.6; 84.0) | 74.0 (64.5; 84.3) | 0.221 |
| Height (cm) | 165.6 ± 6.9 | 179.6 ± 7.1 | <0.001 | 168.7 ± 9.5 | 173.9 ± 9.6 | <0.001 |
| Body mass index (kg/m<sup>2</sup>) | 23.9 (21.7; 27.2) | 25.5 (23.4; 28.4) | <0.001 | 25.5 (23.1; 28.5) | 24.2 (22.1; 26.9) | <0.001 |
| ALM (kg) by BIA | 18.9 ± 2.9 | 27.4 ± 3.8 | <0.001 | 21.1 ± 5.1 | 23.7 ± 5.3 | <0.001 |
| ALM (kg) by DXA | 18.1 ± 2.7 | 26.6 ± 4.0 | <0.001 | 20.3 ± 4.8 | 23.0 ± 5.5 | <0.001 |
| Relative ALM (kg/m<sup>2</sup>) by BIA | 6.8 ± 0.7 | 8.5 ± 0,7 | <0.001 | 7.3 ± 1.1 | 7.7 ± 1.0 | <0.001 |
| Relative ALM (kg/m<sup>2</sup>) by DXA | 6.6 ± 0.8 | 8.2 ± 1 | <0.001 | 7.0 ± 1.1 | 7.5 ± 1.2 | <0.001 |

Values are expressed as median (interquartile range) or mean±SD.

### Appendicular lean mass

There was a strong correlation between the DXA and BIA measurements for both ALM (Figures 1A) and relative ALM (Figure 1B) for all participants. Agreement between BIA and DXA was very high (ICC over 0.90) for ALM for all participants, as well as for men, for women, for younger and older participants, and for relative ALM the ICC was similarly very high for all participants, for younger and older participants, and high (between 0.70 and 0.89) for men and women. (Table 2).

**Figure 1.**
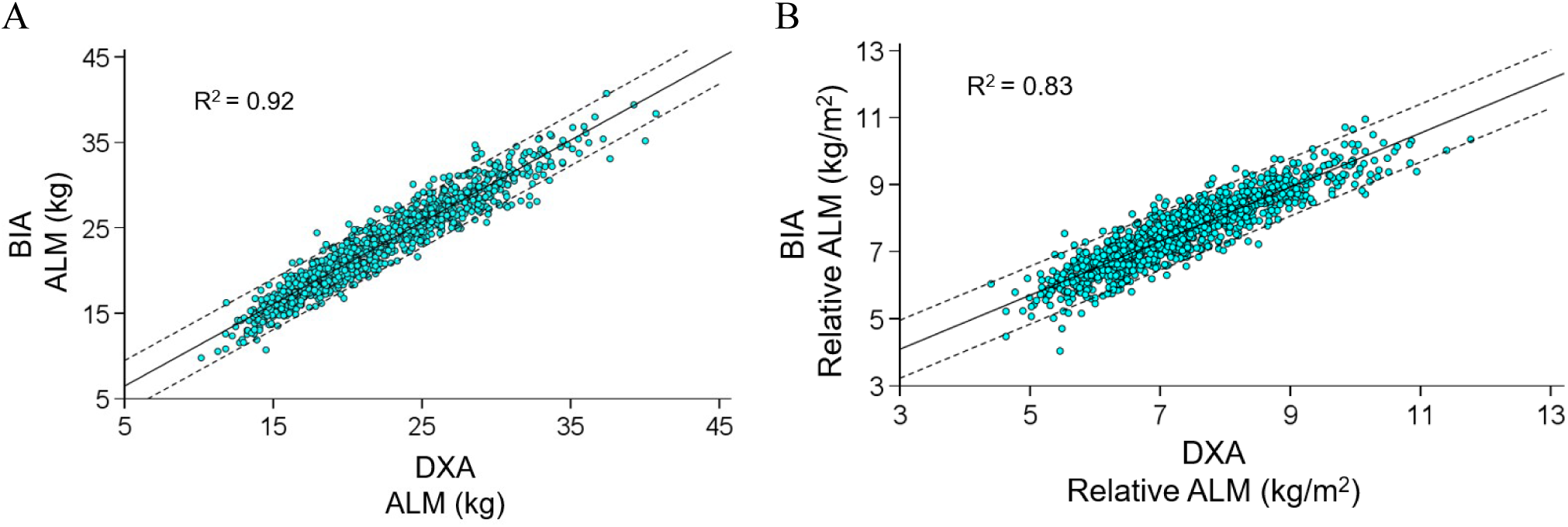
A. Correlation between appendicular lean mass (ALM) by Dual-Energy X-ray absorptiometry (DXA) and Bioimpedance Analysis (BIA) for all participants. B. Correlation between relative ALM by DXA and BIA for all participants. Regression line (solid line) with 95% prediction interval (dashed line) and adjusted R^2^ values are shown.

**Table 2.**
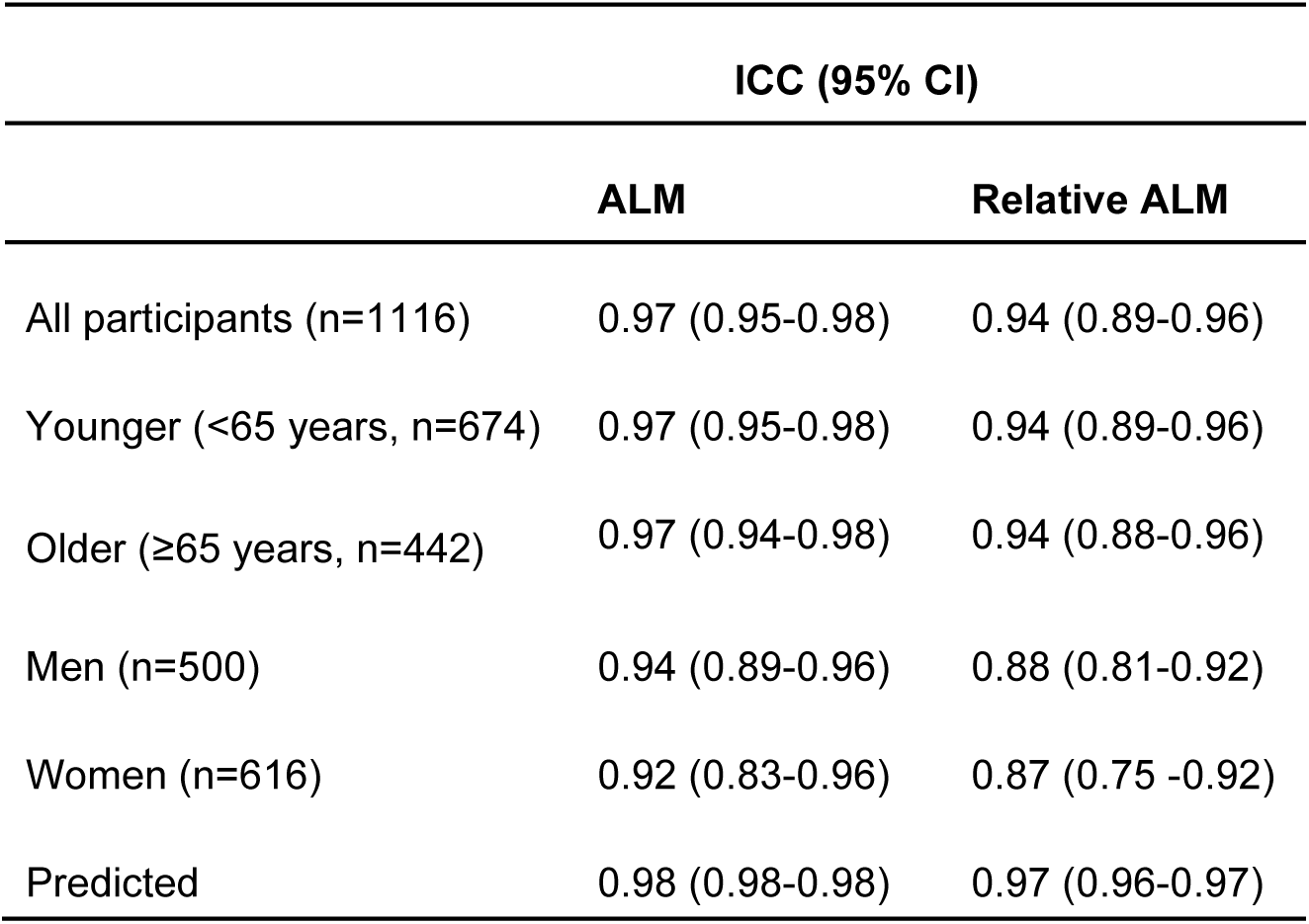
Agreement between BIA and DXA.

Differences between DXA and BIA measures were plotted against the average of the two measurements (Figure 2). The mean difference in ALM for all participants was 0.78 ± 1.53 kg, 0.81 ± 1.77 kg for men, and 0.77 ± 1.31 kg for women. The mean difference in relative ALM was 0.25 ± 0.50 kg/m^2^, 0.24 ± 0.54 kg/m^2^ and 0.26 ± 0.46 kg/m^2^ for all participants, men, and women, respectively. The Bland-Altman plot demonstrated a narrow limit of agreement for ALM and relative ALM measurements, and a systematic small positive bias with an overall overestimation of ALM and relative ALM measurements by BIA.

**Figure 2.**
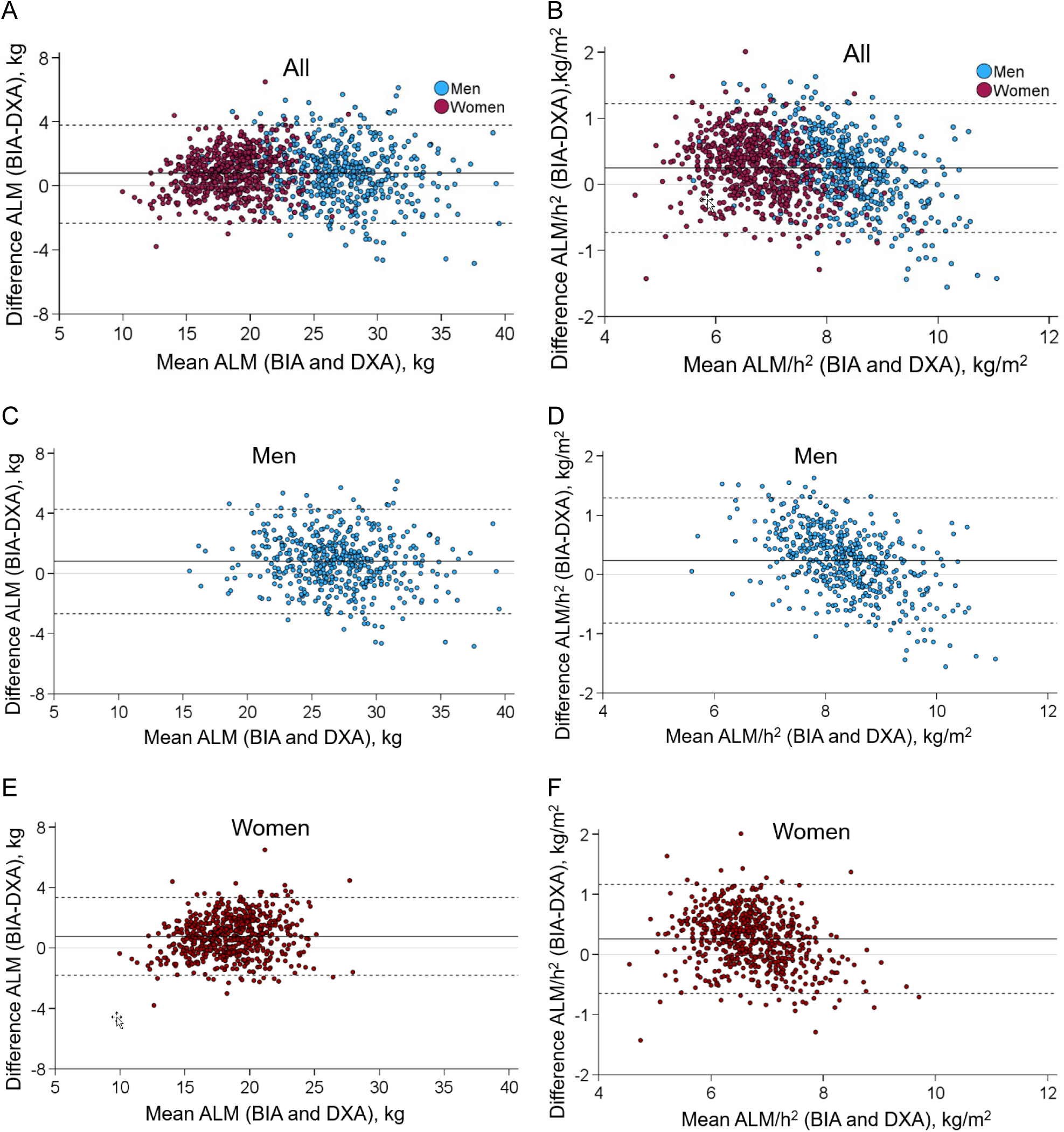
Bland–Altman plot comparing appendicular lean mass (ALM) (A, C and E) and relative ALM (B, D and F) measured by Bioimpedance Analyses (BIA) and Dual-Energy X- ray absorptiometry (DXA). The solid line represents the mean difference. Top and bottom reference lines indicate the limits of agreement (dashed lines).

All participants were divided into two age groups: younger adults (18-64 years, n=674) and older adults (65 years and older, n= 442). Agreements between the two techniques for the two age groups were assessed using Bland-Altman plots (Figure 3). The mean difference in ALM for older adults was 0.81 ± 1.46 kg, and 0.77 ± 1.58 kg, for younger adults while the mean difference in relative ALM was 0.26 ± 0.49 kg/m^2^ and 0.24 ± 0.50 kg/m^2^ respectively, with a tendency for BIA to overestimate ALM and relative ALM. For both groups, there was a narrow limit of agreement on Bland-Altman for ALM and relative ALM measurements, and a systematic small positive bias with an overall overestimation of ALM and relative ALM measurements by BIA. Proportional bias was noted for relative ALM measurement for younger adults, with overestimation by BIA at lower relative ALM and underestimation at higher relative ALM (R^2^=0.2).

**Figure 3.**
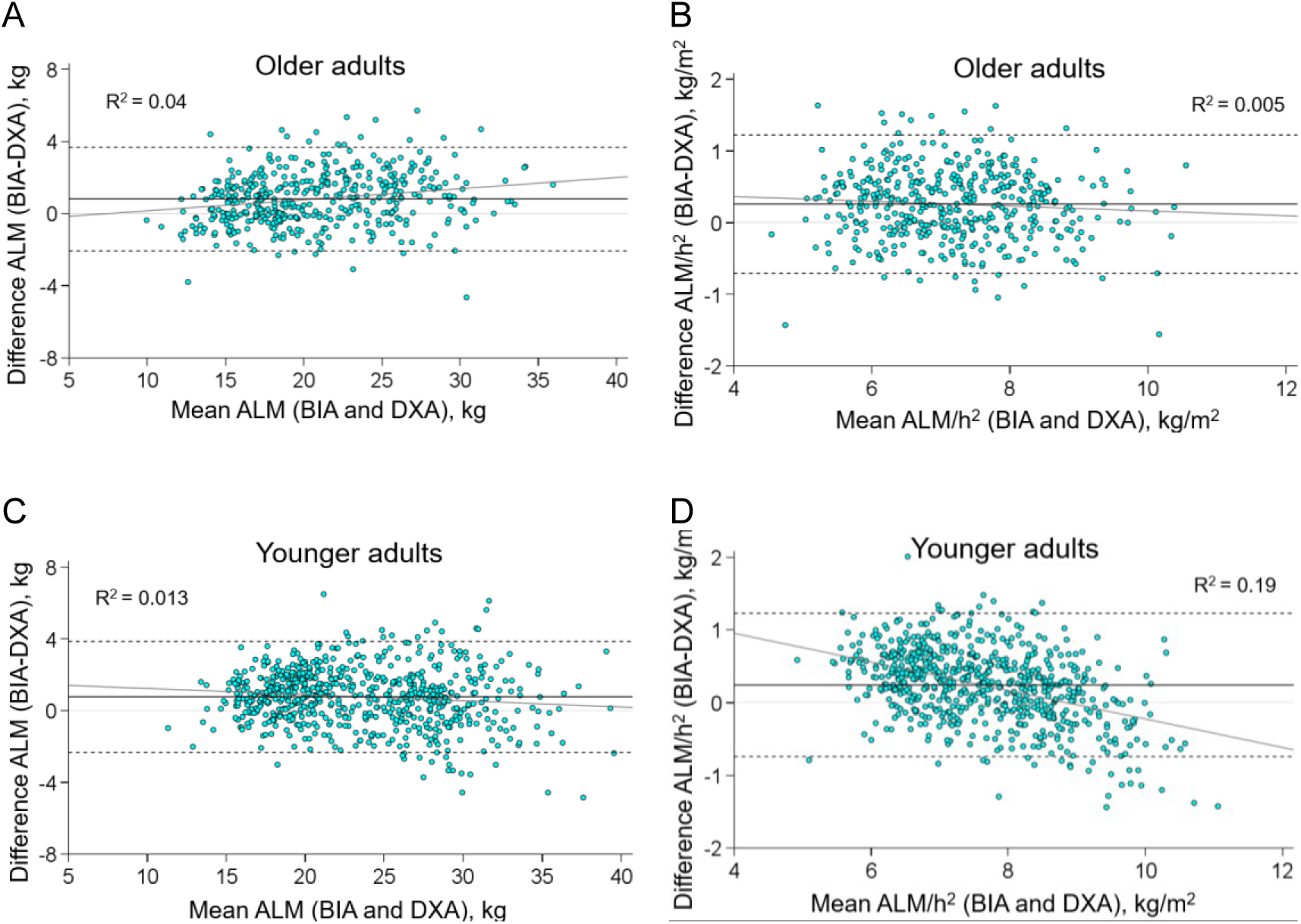
Bland–Altman plot comparing appendicular lean mass (ALM) and relative ALM measured by Bioimpedance Analysis (BIA) and Dual-Energy X-ray absorptiometry (DXA) for older adults (≥65 years) (A, B) and younger adults (<65 years) (C, D). The solid line represents the mean difference. Top and bottom reference lines indicate the limits of agreement (dashed lines).

Linear regression between the measurements obtained by BIA and DXA showed good correlation for ALM (R^2^=0.92). Multivariable regression models included the additional variables BMI, age, height, sex, and weight. The single variable that significantly improved the correlation was the height (p<0.001) explaining 95% of the variance.

The regression analyses demonstrated similar results for relative ALM. Linear regression between the measurements obtained by BIA and DXA showed a good correlation (R^2^=0.83) where the addition of height as a variable improved the correlation significantly (R^2^=0.88) (p<0.001). These results suggest that height correction improves the accuracy of BIA measurements. Indeed, the percent of error for ALM and relative ALM were plotted for different height groups (Figure 4). Measures from BIA tended to slightly underestimate the ALM and relative ALM of participants with a height under 155 cm as opposed to slightly overestimating participants higher than 155 cm.

**Figure 4.**
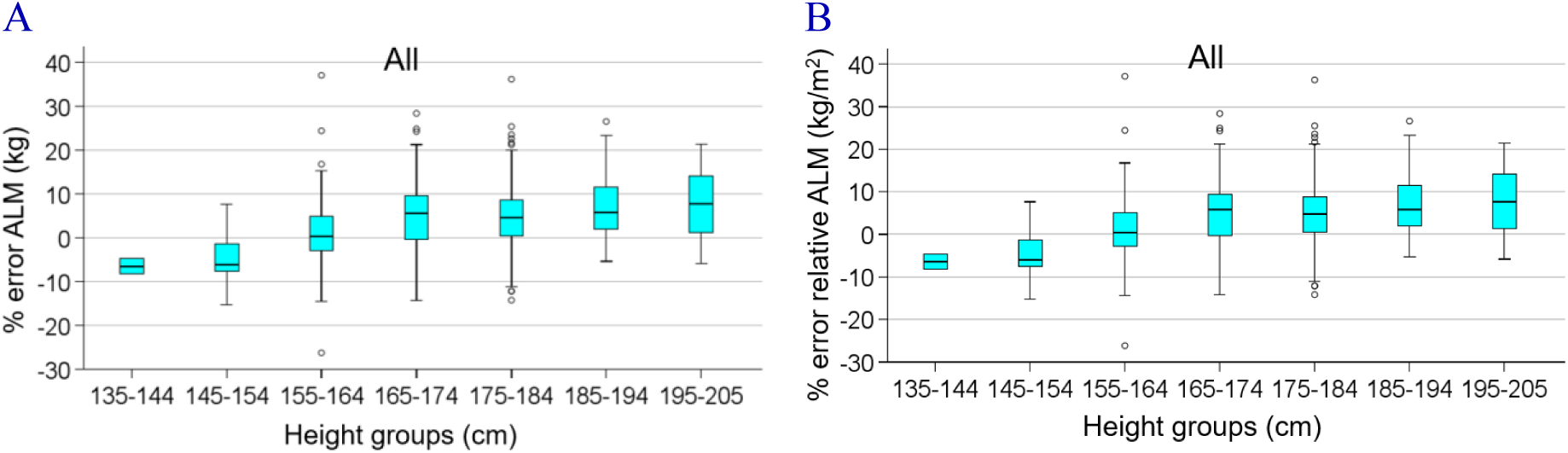
The association between percent of error appendicular lean mass (ALM) and relative ALM at different height groups.

To increase the clinical utility, linear regression equations were formulated to correct the overestimations of BIA compared to DXA. The following equations was developed to achieve an ALM and relative ALM by BIA close to that measured by DXA:

#### For all participants

ALM (DXA) = 27.55 + 1.30 · ALM (BIA) – 0.204 · height (p < 0.001; R^2^ = 0.95). ALM·h^-2^ (DXA) = 4.30 + 1.29 · ALM·h^-2^ (BIA) – 0.039 · height (p < 0.001; R^2^ = 0.88).

#### For older adults

ALM (DXA) = 22.89 + 1.17 · ALM (BIA) – 0.162 · height (p < 0.001; R^2^ = 0.94). ALM·h^-2^ (DXA) = 5.03 + 1.17 · ALM·h^-2^ (BIA) – 0.039 · height (p < 0.001; R^2^ = 0.87).

#### For younger adults

ALM (DXA) = 31.78 + 1.38 · ALM (BIA) – 0.239 · height (p < 0.001; R^2^ = 0.95). ALM·h^-2^ (DXA) = 4.13 + 1.38 · ALM·h^-2^ (BIA) – 0.042 · height (p < 0.001; R^2^ = 0.90).

Differences between the DXA-observed and BIA-predicted ALM and relative ALM were plotted against the average of the two measurements in Bland-Altman plots. The mean difference in ALM for all participants and differentiated for older adults and younger adults was 0.05 ± 1.25 kg, −0.07 ± 1.19 kg and −0.04 ± 1.23 kg, respectively. The mean difference in relative ALM was 0.03 ± 0.42 kg/m^2^, −0.07 ± 0.41 kg/m^2^ and 0.007 ± 0.40 kg/m^2^, for the total group, for older adults, and for younger adults, respectively. Agreement between the DXA-observed and BIA-predicted ALM and relative ALM with low residuals and little evidence of bias (Table 2).

## Discussion

The present study evaluated the agreement between BIA and DXA for measuring ALM and relative ALM in a large, diverse cohort of healthy adults. Our findings demonstrate good agreement between the two methods for the estimation of ALM and relative ALM for all age groups and both sexes. However, consistent with previous studies [22,24], we found that BIA tends to systematically overestimate lean mass by a small margin. Other studies have reported similar biases, particularly in older adults and individuals with a higher BMI [22,24]. However, the present study demonstrated a better limit of agreement than many previous studies and isolated the height of the individual as a contributing factor to the variation between results. Looking closer at the bias, a slight linear trend (R^2^=0.19) was observed within the subgroup of difference of younger adults aged <65 years (22-64 years). Within this group, an overestimation of relative ALM measurements by BIA was demonstrated in subjects having lower relative ALM while descending to an underestimation in subjects having a higher relative ALM. This trend was not observed in the older age-group (≥65 years), which may be partly explained by the lower height variability among subjects in the older group. In contrast to previous studies, however, we found that the agreement between ALM and relative ALM obtained by BIA and DXA is very high with ICC’s of 0.97 and 0.94, respectively. Compared to DXA, we found BIA to overestimate ALM by an average of 0.78 kg and relative ALM by 0.25 kg/m^2^, which is remarkably less than observed previously published studies [20,22,23,29].

Furthermore, regression equations for the BIA-derived estimate of ALM and relative ALM were developed to provide the possibility to convert BIA derived measurements to values even more concordant with DXA. We found height to be the variable that best accounted for differences in BIA-derived ALM and relative ALM and those from DXA. For this reason, we propose a formula to produce corrected BIA ALM measures with accounts for height, which corrects 95% of the deviation from DXA ALM measures. Likewise, we present a formula that improves the BIA relative ALM estimates by again correcting for height, which explains 88% of the variation. We have not been able to find similar equations in previous studies that have developed formulas using sex and BMI or other parameters [20,22,29].

The improved agreement between BIA and DXA ALM measurements in this study can partially be attributed to the model of the BIA device used. Several studies have highlighted the importance of device-specific variability in BIA measurements. For instance, Reiss et al. (2016) emphasized that different BIA devices produce varying results, even when compared to the same DXA reference [23]. Our study focused on the InBody 770, a multi-frequency BIA device that is commonly used in clinical settings [30–32]. While our results demonstrate strong agreement between BIA and DXA, it is important to recognize that these findings may not be generalizable to all BIA devices. Beyond studies such as this being device specific, the size and type of cohort chosen is important [20,29] and can be limiting for the application of the study’s results to a broad population [21–23]. For this reason, the sample size of subjects in the present study is comparatively large and diverse in age, sex, and body composition, but not ethnicity.

### Clinical Implications

The findings of this study have significant clinical implications, particularly for the assessment of lean mass and, thereby, sarcopenia in older adults. Early detection of sarcopenia is critical for implementing interventions aimed at preserving muscle mass and function. Given the portability, affordability, and ease of use of BIA, it is a valuable tool for clinicians who need to assess lean mass in settings where DXA is not available. The correction models developed in this study provide a practical means of adjusting BIA measurements to align more closely with DXA, making BIA a more reliable option for routine clinical use.

The overestimation of lean mass by BIA, particularly in younger adults, may reflect differences in muscle quality and hydration status that BIA cannot fully capture. Therefore, while BIA can serve as a useful screening tool, it should be used with caution in populations with high muscle mass or abnormal body composition.

### Limitations

While this study provides valuable insights, it has several limitations. The study population consisted of healthy individuals, which may limit the generalizability of the findings to populations with chronic diseases or conditions that affect body composition, such as cancer, heart failure, or obesity.

Furthermore, while the InBody 770 is a widely used BIA device, our findings may not be fully applicable to other BIA devices, which have different algorithms and measurement techniques. Future studies should focus on validating these results across different BIA devices and in populations with varying health statuses.

### Future Directions

Future research should aim to validate the correction models developed in this study in larger and more diverse populations, including those with chronic illnesses and significant body composition abnormalities. Longitudinal studies are needed to assess the utility of BIA in tracking lean mass changes over time and predicting clinical outcomes. Additionally, further investigation into the effects of hydration status, fat distribution, and ethnic differences on BIA accuracy would enhance our understanding of how to optimize BIA for use in diverse clinical settings.

### Conclusion

In conclusion, this study demonstrates that BIA is a reliable method for estimating appendicular lean mass (ALM) and relative ALM, with strong agreement with DXA. However, BIA tends to overestimate lean mass, particularly in younger adults. By applying the regression equations developed in this study, clinicians can obtain BIA measurements that are more accurate, which improves BIA as a practical alternative to DXA in clinical settings where DXA is not accessible. These findings are especially relevant for the early detection and management of sarcopenia in older adults, allowing for timely interventions to preserve muscle function and reduce the risk of adverse health outcomes. Further research is necessary to validate these models in more diverse populations and explore the long-term clinical utility of BIA in assessing lean mass.

## Data Availability

All data produced in the present work are contained in the manuscript

## References

1 Aversa Z, Zhang X, Fielding RA, et al. The clinical impact and biological mechanisms of skeletal muscle aging. Bone 2019;127:26–36. doi:10.1016/j.bone.2019.05.021

2 McLeod M, Breen L, Hamilton DL, et al. Live strong and prosper: the importance of skeletal muscle strength for healthy ageing. Biogerontology 2016;17:497–510. doi:10.1007/s10522-015-9631-7

3 Gandham A, Gregori G, Johansson L, et al. Sarcopenia definitions and their association with injurious falls in older Swedish women from the Sahlgrenska University Hospital Prospective Evaluation of Risk of Bone fractures (SUPERB) study. Osteoporos Int 2024;35:1963–72. doi:10.1007/s00198-024-07196-0

4 Gandham A, Gregori G, Johansson L, et al. Sarcopenia definitions and their association with fracture risk in older Swedish women. J Bone Miner Res 2024;39:453–61. doi:10.1093/jbmr/zjae026

5 Thackeray M, Mohebbi M, Orford N, et al. Lean mass as a risk factor for intensive care unit admission: an observational study. Crit Care 2021;25:364. doi:10.1186/s13054-021-03788-y

6 Xu J, Wan CS, Ktoris K, et al. Sarcopenia Is Associated with Mortality in Adults: A Systematic Review and Meta-Analysis. Gerontology 2022;68:361–76. doi:10.1159/000517099

7 Beaudart C, Zaaria M, Pasleau F, et al. Health Outcomes of Sarcopenia: A Systematic Review and Meta-Analysis. PLoS ONE 2017;12:e0169548. doi:10.1371/journal.pone.0169548

8 Rivera FB, Escolano BT, Nifas FM, et al. Interrelationship of sarcopenia and cardiovascular diseases: A review of potential mechanisms and management. JAFES 2024;39:69–78. doi:10.15605/jafes.039.01.03

9 Kang S-Y, Lim GE, Kim YK, et al. Association between Sarcopenic Obesity and Metabolic Syndrome in Postmenopausal Women: A Cross-sectional Study Based on the Korean National Health and Nutritional Examination Surveys from 2008 to 2011. J Bone Metab 2017;24:9–14. doi:10.11005/jbm.2017.24.1.9

10 Izzo A, Massimino E, Riccardi G, et al. A narrative review on sarcopenia in type 2 diabetes mellitus: prevalence and associated factors. Nutrients 2021;13. doi:10.3390/nu13010183

11 Stone L, Olson B, Mowery A, et al. Association between sarcopenia and mortality in patients undergoing surgical excision of head and neck cancer. JAMA Otolaryngol Head Neck Surg 2019;145:647–54. doi:10.1001/jamaoto.2019.1185

12 Brown JC, Schmitz KH. Weight lifting and appendicular skeletal muscle mass among breast cancer survivors: a randomized controlled trial. Breast Cancer Res Treat 2015;151:385–92. doi:10.1007/s10549-015-3409-0

13 McGregor RA, Cameron-Smith D, Poppitt SD. It is not just muscle mass: a review of muscle quality, composition and metabolism during ageing as determinants of muscle function and mobility in later life. Longev Healthspan 2014;3:9. doi:10.1186/2046-2395-3-9

14 Beaudart C, McCloskey E, Bruyère O, et al. Sarcopenia in daily practice: assessment and management. BMC Geriatr 2016;16:170. doi:10.1186/s12877-016-0349-4

15 Cruz-Jentoft AJ, Baeyens JP, Bauer JM, et al. Sarcopenia: European consensus on definition and diagnosis: Report of the European Working Group on Sarcopenia in Older People. Age Ageing 2010;39:412–23. doi:10.1093/ageing/afq034

16 Buckinx F, Landi F, Cesari M, et al. Pitfalls in the measurement of muscle mass: a need for a reference standard. J Cachexia Sarcopenia Muscle 2018;9:269–78. doi:10.1002/jcsm.12268

17 Erlandson MC, Lorbergs AL, Mathur S, et al. Muscle analysis using pQCT, DXA and MRI. Eur J Radiol 2016;85:1505–11. doi:10.1016/j.ejrad.2016.03.001

18 Cruz-Jentoft AJ, Bahat G, Bauer J, et al. Sarcopenia: revised European consensus on definition and diagnosis. Age Ageing 2019;48:16–31. doi:10.1093/ageing/afy169

19 Yamada Y, Nishizawa M, Uchiyama T, et al. Developing and Validating an Age-Independent Equation Using Multi-Frequency Bioelectrical Impedance Analysis for Estimation of Appendicular Skeletal Muscle Mass and Establishing a Cutoff for Sarcopenia. Int J Environ Res Public Health 2017;14. doi:10.3390/ijerph14070809

20 Yamada Y, Yamada M, Yoshida T, et al. Validating muscle mass cutoffs of four international sarcopenia-working groups in Japanese people using DXA and BIA. J Cachexia Sarcopenia Muscle 2021;12:1000–10. doi:10.1002/jcsm.12732

21 Scafoglieri A, Clarys JP, Bauer JM, et al. Predicting appendicular lean and fat mass with bioelectrical impedance analysis in older adults with physical function decline - The PROVIDE study. Clin Nutr 2017;36:869–75. doi:10.1016/j.clnu.2016.04.026

22 Buckinx F, Reginster J-Y, Dardenne N, et al. Concordance between muscle mass assessed by bioelectrical impedance analysis and by dual energy X-ray absorptiometry: a cross-sectional study. BMC Musculoskelet Disord 2015;16:60. doi:10.1186/s12891-015-0510-9

23 Reiss J, Iglseder B, Kreutzer M, et al. Case finding for sarcopenia in geriatric inpatients: performance of bioimpedance analysis in comparison to dual X-ray absorptiometry. BMC Geriatr 2016;16:52. doi:10.1186/s12877-016-0228-z

24 Kim JH, Choi SH, Lim S, et al. Assessment of appendicular skeletal muscle mass by bioimpedance in older community-dwelling Korean adults. Arch Gerontol Geriatr 2014;58:303–7. doi:10.1016/j.archger.2013.11.002

25 Suetta C, Haddock B, Alcazar J, et al. The Copenhagen Sarcopenia Study: lean mass, strength, power, and physical function in a Danish cohort aged 20-93 years. J Cachexia Sarcopenia Muscle 2019;10:1316–29. doi:10.1002/jcsm.12477

26 Baarts RB, Jensen MR, Hansen OM, et al. Age- and sex-specific changes in visceral fat mass throughout the life-span. Obesity (Silver Spring) 2023;31:1953–61. doi:10.1002/oby.23779

27 Bland JM, Altman DG. Measuring agreement in method comparison studies. Stat Methods Med Res 1999;8:135–60. doi:10.1177/096228029900800204

28 Koo TK, Li MY. A guideline of selecting and reporting intraclass correlation coefficients for reliability research. J Chiropr Med 2016;15:155–63. doi:10.1016/j.jcm.2016.02.012

29 Cheng KY-K, Chow SK-H, Hung VW-Y, et al. Diagnosis of sarcopenia by evaluating skeletal muscle mass by adjusted bioimpedance analysis validated with dual-energy X-ray absorptiometry. J Cachexia Sarcopenia Muscle 2021;12:2163–73. doi:10.1002/jcsm.12825

30 Falbová D, Beňuš R, Sulis S, et al. Effect of COVID-19 pandemic on bioimpedance health indicators in young adults. Am J Hum Biol 2024;36:e24110. doi:10.1002/ajhb.24110

31 Huang S, Chen X, Ding H, et al. The relationship between low and asymmetric handgrip strength and low muscle mass: results of a cross-sectional study on health and aging trends in western China. BMC Geriatr 2024;24:650. doi:10.1186/s12877-024-05199-4

32 McLester CN, Nickerson BS, Kliszczewicz BM, et al. Reliability and Agreement of Various InBody Body Composition Analyzers as Compared to Dual-Energy X-Ray Absorptiometry in Healthy Men and Women. J Clin Densitom 2020;23:443–50. doi:10.1016/j.jocd.2018.10.008

